# Trends in Antibiotic Use among Cardiovascular Heart Disease Inpatients at the Jakaya Kikwete Cardiac Institute in Tanzania from 2016 to 2022

**DOI:** 10.1101/2025.09.24.25336595

**Authors:** Jackob Aron Ndayomwami, Judith Ambele Mwamelo, Hamis Abdalla Kaniki, James Mwakyomo, Naizihijwa Majani, Reuben Kato Mutagaywa, Peter Kisenge, Mohamed Janabi, Raphael Z. Sangeda

## Abstract

**Objective:** To analyse trends in antibiotic use among cardiovascular inpatients at the Jakaya Kikwete Cardiac Institute (JKCI), Tanzania, from 2016 to 2022, using the World Health Organisation (WHO) Anatomical Therapeutic Chemical/Defined Daily Dose (ATC/DDD) methodology.

**Design:** Retrospective longitudinal study.

**Setting:** Jakaya Kikwete Cardiac Institute, a 150-bed national referral and teaching hospital in Dar es Salaam, Tanzania.

**Patients:** 11,656 cardiovascular inpatients; 6,612 (56.7%) received at least one systemic antibiotic.

**Methods:** Prescription data (250,591 total; 30,885 antibiotic-related) were extracted from the JKCI MedPro dispensing system. Antibiotic use was expressed in Defined Daily Doses per 100 bed-days (DDD/100). Analyses were conducted using Microsoft Excel, SPSS version 26, and time-series forecasting with AutoRegressive Integrated Moving Average (ARIMA) modelling in R.

**Results:** Overall, antibiotic use was 29.88 DDD/100 bed-days. Injections accounted for more than 55% of use. At the ATC Class 4 level, carbapenems (J01DH) dominated (45.6%), followed by third-generation cephalosporins (17.0%) and extended-spectrum penicillins (14.6%). At the molecular level, meropenem contributed 45.6%, with amoxicillin and ceftriaxone also being prominent; the top 11 antibiotics comprised more than 90% of total use (Drug Utilisation 90% [DU90%]). By WHO Access, Watch, Reserve (AWaRe) classification, Reserve agents contributed 46.4% of use, followed by Watch (31.3%) and Access (21.1%). ARIMA (1,1,0) modelling forecasts a continued upward trend through 2026.

**Conclusions:** Antibiotic use among cardiovascular inpatients at JKCI is high, with heavy reliance on broad-spectrum and Reserve agents. Strengthened antimicrobial stewardship measures, such as intravenous-to-oral switch protocols, DU90 audits, and stricter control of Reserve antibiotic prescribing, are urgently needed to mitigate antimicrobial resistance risks.

## 1. Introduction

Cardiovascular diseases (CVDs)—a group that includes coronary heart disease (CHD), heart attacks, arrhythmias, heart failure, and cardiomyopathy—remain the leading cause of death globally ^1–3^. CHD alone is responsible for approximately 16% of all global deaths and contributes significantly to the need for cardiac interventions and surgeries. Recent estimates from the Global Burden of Disease Study indicate that CVDs accounted for 19.8 million deaths in 2022, up from 12.4 million deaths in 1990, marking a 59.7% increase in absolute terms ^4,5^. Despite this rise in absolute mortality, the age-standardised death rate from CVDs has declined by 34.9% over the same period, suggesting improvements in preventive measures and clinical care.

Patients with CVD, particularly the elderly, frequently receive antibiotics due to their increased susceptibility to bacterial infections. Although early studies suggested that antibiotic use might reduce the risk of CHD by mitigating chronic infections and systemic inflammation ^6^, subsequent evidence has challenged this hypothesis. Long-term antibiotic exposure—especially macrolides such as azithromycin—has been associated with an elevated risk of cardiovascular events in older women ^7^. Additionally, antibiotics may disrupt the host microbiota, contributing to increased risks of stroke and heart disease ^8^.

Antimicrobial resistance (AMR), now recognised by the WHO as one of the top ten global health threats, further complicates the use of antibiotics in high-risk populations. AMR occurs when pathogens evolve to survive exposure to antimicrobial agents, rendering standard treatments ineffective ^9^. This resistance is largely driven by the indiscriminate use of antibiotics in both human and veterinary medicine. Without targeted interventions, AMR could lead to as many as 10 million deaths annually by 2050 ^10^.

The burden of AMR is especially severe in low- and middle-income countries (LMICs), where prolonged illness, limited access to second-line treatments, and higher mortality rates present significant public health challenges. Between 2000 and 2010, global antibiotic consumption rose by 36%, with Brazil, Russia, India, China, and South Africa accounting for approximately 76% of this increase ^11^. In LMICs, increasing antibiotic access due to economic development has been accompanied by misuse, influenced by cultural prescribing norms, limited diagnostic capacity, and weak regulatory enforcement ^12,13^. Vulnerable populations—particularly children and the elderly—frequently receive antibiotics due to age-related immune vulnerabilities ^14–16^.

While Point Prevalence Surveys (PPS) have traditionally been used to assess antibiotic use in hospital settings, they offer only cross-sectional insights. In contrast, the WHO recommends the use of Defined Daily Dose (DDD) per 100 bed-days for longitudinal surveillance in inpatient populations ^17^. This approach enables more accurate monitoring of antibiotic use trends and evaluation of antimicrobial stewardship (AMS) interventions in high-acuity environments.

In Tanzania, there is limited facility-level data on inpatient antibiotic use. WHO AMR surveillance reports from 2015 to 2016 showed that only 4 of 54 African countries submitted consumption data. Tanzania reported a rate of 27.29 DDD/1000 inhabitants/day, while Burundi recorded the lowest at 4.44 ^18^. National antibiotic consumption from 2017 to 2019 averaged 1.5 million DDDs per year, translating to 80.8 DDDs per 1,000 inhabitants per day. 19,20 However, these national estimates do not capture institution-specific or disease-specific trends, particularly in specialised populations, such as those with cardiovascular conditions.

This study aimed to address this gap by analysing antibiotic use among cardiovascular inpatients at the Jakaya Kikwete Cardiac Institute (JKCI), Tanzania’s national referral centre for cardiac care. We employed the WHO ATC/DDD methodology. We used bed-days as the denominator to evaluate longitudinal trends in antibiotic use from 2016 to 2022 and explore their implications for AMS in specialised cardiac settings.

## 2. Materials and Methods

### Study Design and Approach

This was a retrospective longitudinal study that examined trends in antibiotic use among CVD inpatients over a six-year period (July 2016 – June 2022) at the JKCI in Dar es Salaam, Tanzania. Antibiotic use was assessed using the Anatomical Therapeutic Chemical (ATC) / DDD methodology established by the World Health Organisation (WHO, 2003), and results were expressed in DDD per 100 bed-days to standardise comparisons across time and patient categories.

### Study Setting

JKCI is a national referral hospital specialising in cardiovascular care. It is also a university teaching and research facility that was established in 2015. The Institute has a 150-bed capacity and manages around 100 inpatients and 700 outpatients per week. Services range from elective procedures to emergency and intensive cardiac care, making it a high-acuity environment where antibiotics are frequently used for prophylaxis and treatment. It is located in Dar es Salaam, Tanzania, within the Ilala District in Upanga West ward.

### Study Population and Data Sources

During the six-year study period, a total of 250,591 prescriptions were issued to 11,656 patients with cardiovascular conditions. Of these, 30,885 prescriptions (12.3%) were related to antibiotics, and 6,612 patients (56.7%) received at least one systemic antibiotic. These data were retrieved from MedPro, JKCI’s electronic pharmacy dispensing system, which integrates medication records from both inpatient and cardiology pharmacies.

Each prescription record extracted from the MedPro system contained essential variables required for DDD-based analysis. These included the dispensing date, the brand and generic names of the antibiotic, the dosage form and strength, the total quantity dispensed and patient demographics such as age and gender. Additionally, admission and discharge dates were recorded to calculate the length of stay for inpatients and the number of bed days for each patient.

### Inclusion and exclusion criteria

Only systemic antibiotics (oral and parenteral) prescribed to inpatients were included in the analysis. **Non-systemic antibiotics** such as topical, ophthalmic, otic, and other local applications were **excluded**. Outpatient antibiotic prescriptions were also excluded from the study.

### Data Management and Analysis

Data cleaning, transformation, and calculation of antibiotic use metrics were conducted using Microsoft Excel (2013) with Power Query. Additional patient-level summary statistics were generated using SPSS Version 26.

DDD and DDD per 100 bed days were calculated in Power BI using the following formulas ^21^.

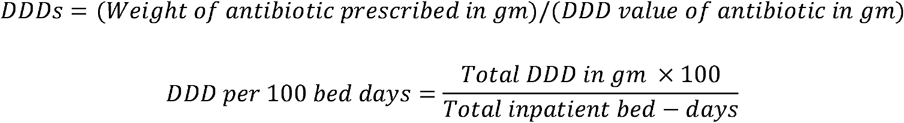

For this study, a bed-day was defined as a full 24-hour hospital stay by a single patient. Total inpatient stay durations were calculated using admission and discharge dates. Antibiotics were further classified using the 2021 WHO Access, Watch, and Reserve (AWaRe) framework, which designates Access agents for common infections, Watch agents for those with higher resistance risk, and Reserve agents as last-line therapies. In addition, Drug Utilisation 90% (DU90) was calculated by ranking antibiotics according to their volume of use and identifying the number of agents that account for 90% of the total Defined Daily Doses (DDD).

### Forecasting Antibiotic Use Trends

To evaluate annual trends in inpatient antibiotic use at JKCI, time-series analysis was applied to Defined Daily Doses (DDD) per 100 bed-days from fiscal years 2016–2017 through 2021– 2022. Forecasting was performed using an AutoRegressive Integrated Moving Average (ARIMA) model in R (forecast package). Model selection was guided by autocorrelation and partial autocorrelation diagnostics, residual patterns, and Akaike Information Criterion (AIC). The best-fitting model, ARIMA (1,1,0), was used to project antibiotic use through 2025– 2026, with 95% confidence intervals to account for prediction uncertainty..

### Ethical considerations

The study was approved by the Muhimbili University of Health and Allied Sciences (MUHAS) Research and Ethics Committee under reference number DA.25/111/25/01/2021, in collaboration with JKCI. Patient confidentiality was strictly maintained—no patient names or personal identifiers were used; only anonymised, aggregated data were analysed.

## 3. Results

### Demographic Characteristics of Inpatients

A total of 6,612 inpatients were included in the analysis. The gender distribution was nearly equal, with 50.4% of the population being male. Most patients were insured by the National Health Insurance Fund (NHIF, 64.3%), followed by private schemes (30.3%). Pediatric patients aged 0–9 years comprised 28.7% of admissions, while those aged 60 years or older comprised 34.0%. Admissions were concentrated in fiscal years 2018–2019 through 2021– 2022. Overall, 79.7% of patients completed treatment within a single fiscal year, and 20.3% had admissions spanning multiple years (Table 1).

**Table 1:**
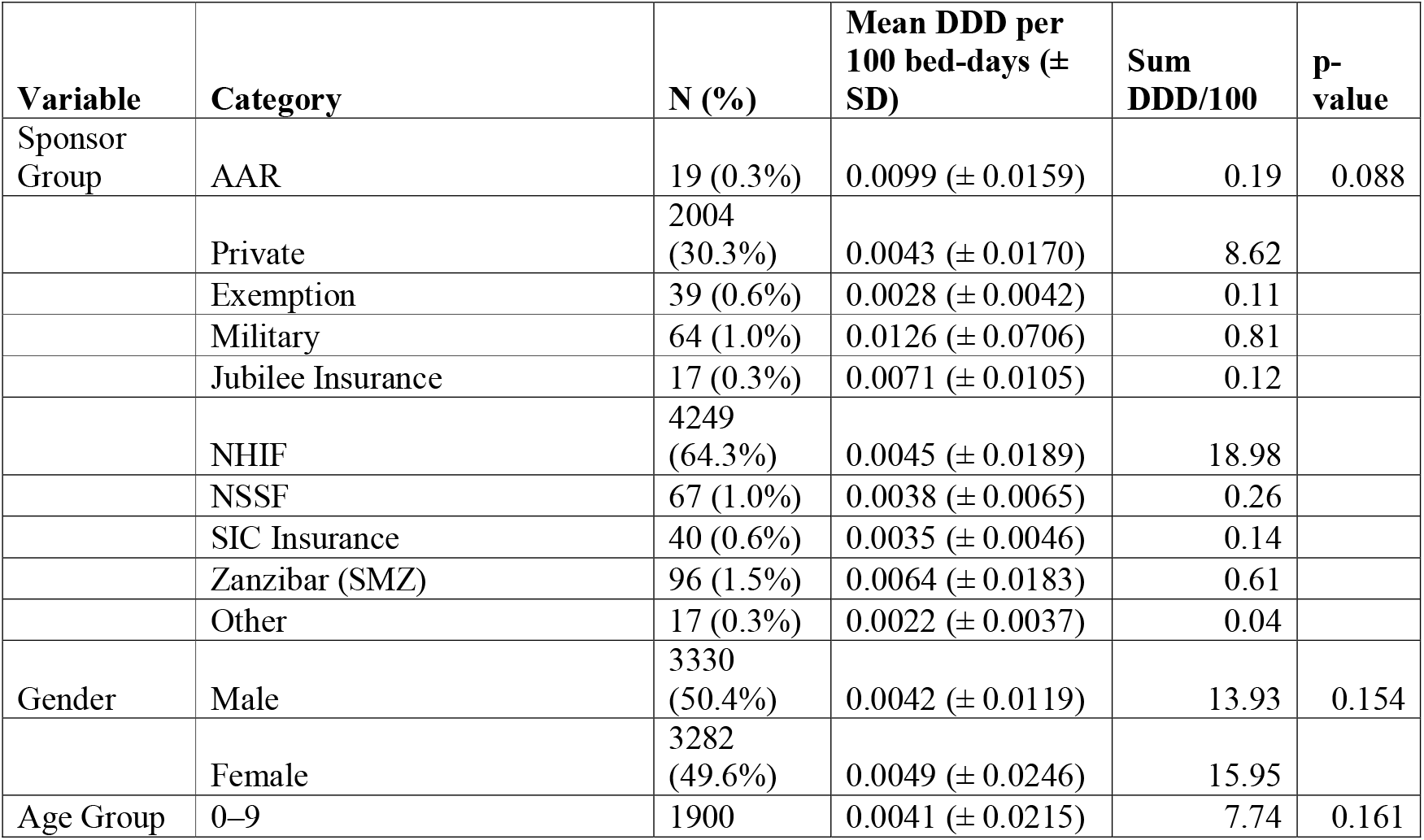
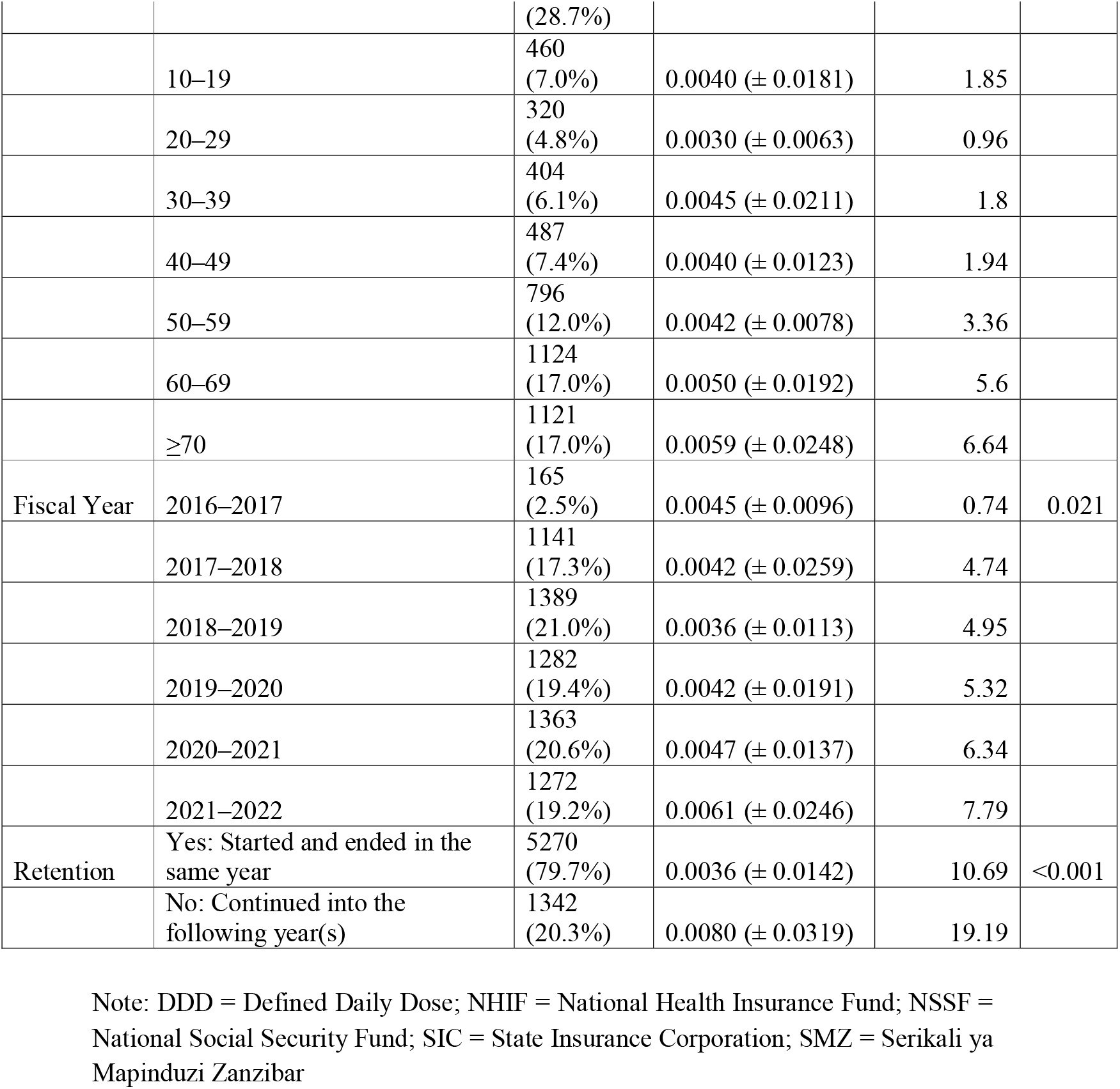
Baseline characteristics of patients in terms of sex and age groups.

### Mean antibiotic use among inpatients

Antibiotic use, expressed as DDD per 100 bed-days, varied across patient and administrative characteristics (Table 1). By fiscal year, mean values increased from 0.0045 in 2016–2017 to 0.0061 in 2021–2022 (p = 0.021). Across age groups, mean use ranged from 0.0030 in patients aged 20–29 years to 0.0059 in those aged 70 years and above, with no statistically significant differences (p = 0.161). Retention status was associated with variation in use (p < 0.001): patients whose admissions extended into subsequent years recorded a mean of 0.0080 DDD/100, compared with 0.0036 among those completing treatment within a single year. No significant differences were observed between genders (p = 0.154) or sponsor groups (p = 0.088). (Table 1).

### Total antibiotic use in inpatients

Overall, antibiotic use among cardiovascular inpatients between 2016 and 2022 was 29.88 Defined Daily Doses (DDD) per 100 bed-days (Table 1). By age group, cumulative use was highest among children aged 0–9 years (7.74 DDD/100) and patients aged 70 years and older (6.64 DDD/100), while the lowest was in the 20–29 years group (0.96 DDD/100). With respect to sponsor categories, patients covered by the National Health Insurance Fund (NHIF) accounted for the largest share of use (18.98 DDD/100), followed by privately sponsored patients (8.62 DDD/100). Annual totals increased over time, rising from 0.74 DDD/100 in 2016–2017 to 7.79 DDD/100 in 2021–2022. By retention status, patients with admissions extending into the following year contributed 19.19 DDD/100, compared with 10.69 among those whose episodes ended within a single fiscal year. (Table 1).

### Antibiotic use in inpatients by dosage form

Antibiotic use among inpatients differed by dosage form (Figure 1). Injections contributed the largest share at 16.57 Defined Daily Doses (DDD) per 100 bed-days, representing 55.4% of total use. Tablets followed with 9.59 DDD/100 (32.1%), while syrups and capsules contributed 2.06 DDD/100 (6.9%) and 1.66 DDD/100 (5.5%), respectively. Infusions were rarely used, accounting for less than 0.01% of total inpatient antibiotic use (0.0004 DDD/100).

**Figure 1:**
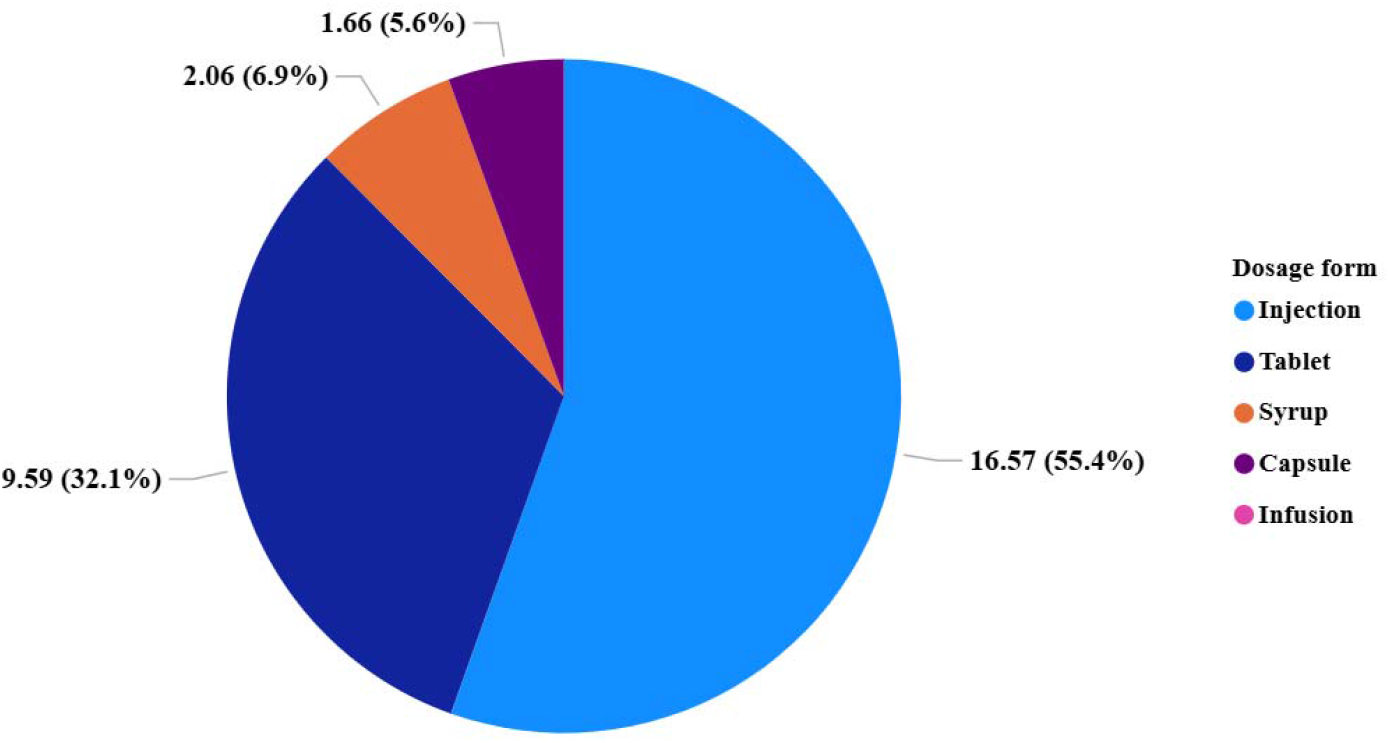
Inpatient antibiotic use by dosage form among cardiovascular heart disease patients at JKCI, 2016–2022 (Daily Defined Doses per 100 bed-days).

### Class-specific trends

Analysis of inpatient antibiotic use by ATC Class 4 groups revealed that carbapenems (J01DH) were the most frequently consumed antibiotic class, contributing 13.63 DDD per 100 bed-days, and accounting for 45.6% of total inpatient antibiotic use across all study years (Figure 2, Panel A). This was followed by third-generation cephalosporins (J01DD) at 5.06 DDD per 100 bed-days (17.0%) and penicillins with extended spectrum (J01CA) at 4.37 DDD per 100 bed-days (14.6%). The remaining antibiotic classes collectively represented less than 23% of total use. Among these, the highest contributions were from macrolides (J01FA), second-generation cephalosporins (J01DC), and imidazole derivatives (J01XD), each contributing between 1.1 and 1.7 DDD per 100 bed-days (Figure 2, Panel B).

**Figure 2:**
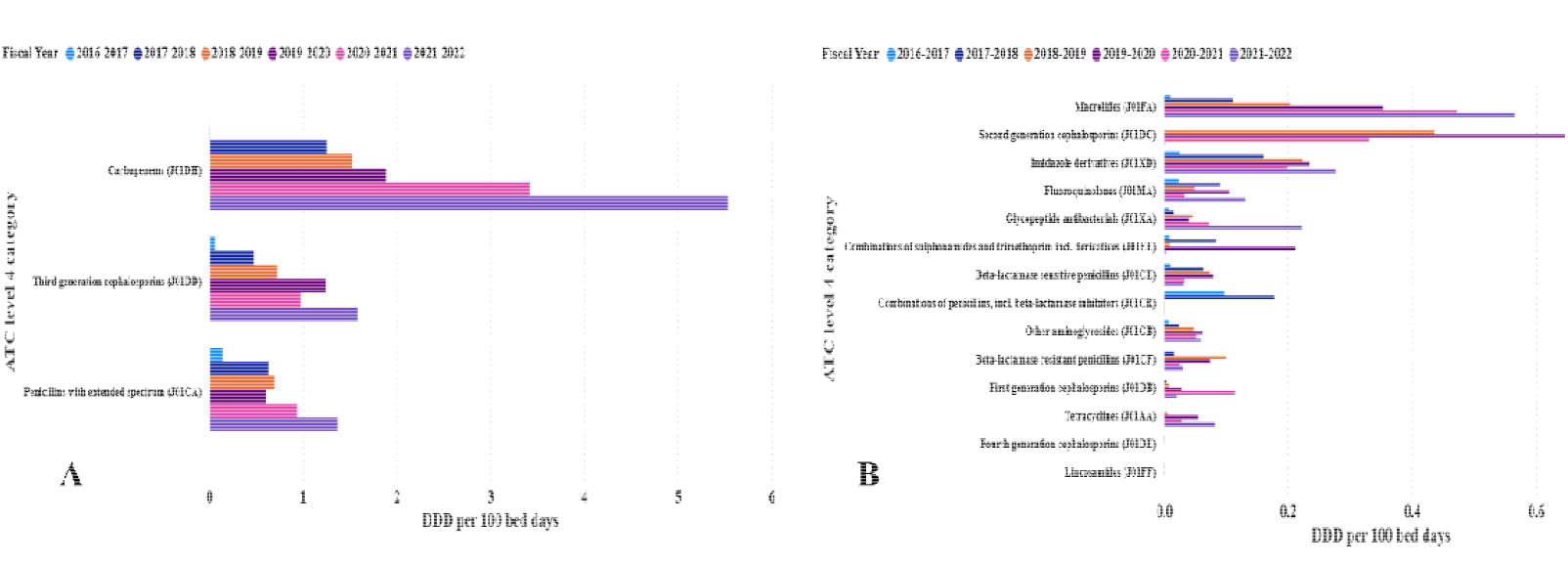
Contribution of ATC Class 4 antibiotics to inpatient use at JKCI, Tanzania, from 2016 to 2022. **Panel A**: Top three most consumed antibiotic classes — carbapenems and **Panel B**: Remaining Anatomical Therapeutic Chemical classification Class 4 antibiotic classes with lower total Defined Daily Dose per 100 bed-days, presented in descending order of use.

Notably, carbapenem use showed a consistent upward trend over the study period, increasing from 0.01 DDD per 100 bed-days in 2016–2017 to 5.54 in 2021–2022—a more than 400-fold rise. This escalation was particularly pronounced from 2019 onwards. Several lower-use classes, such as fourth-generation cephalosporins (J01DE) and lincosamides (J01FF), contributed negligibly, accounting for less than 0.01% of total use.

At the molecule level (ATC Level 5), meropenem (J01DH02) was the most consumed antibiotic, accounting for 13.63 DDD per 100 bed-days, representing 45.6% of all inpatient antibiotic use (Figure 3). Other highly used agents included amoxicillin (J01CA04) at 11.**7%**, ceftriaxone (J01DD04) at 10.9%, and cefuroxime (J01DC02) at 4.7**%**. The top 11 antibiotics cumulatively accounted for over 91**%** of total antibiotic use.

**Figure 3:**
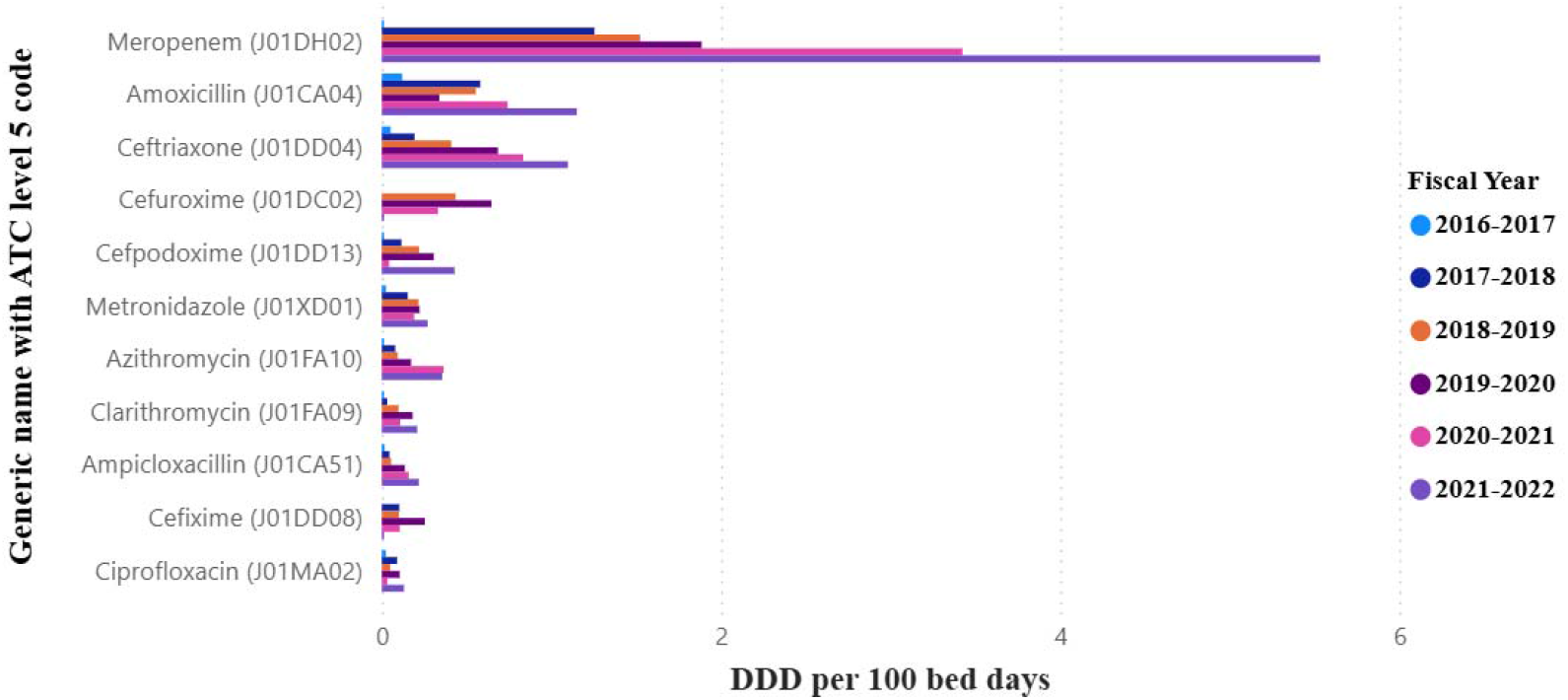
Top 11 most consumed antibiotic molecules (Anatomical Therapeutic Chemical (ATC) classification Level 5) among inpatients at JKCI from 2016 to 2022, shown by Defined Daily Dose (DDD) per 100 bed-days and percentage contribution.

The remaining antibiotics, each contributing less than 1.5% (Table 2), include vancomycin, cotrimoxazole, gentamicin, and doxycycline. Very low-use molecules, including erythromycin, imipenem**-**cilastatin, and cefepime, each accounted for less than 0.01**%** of total DDDs (Table 2).

**Table 2:**
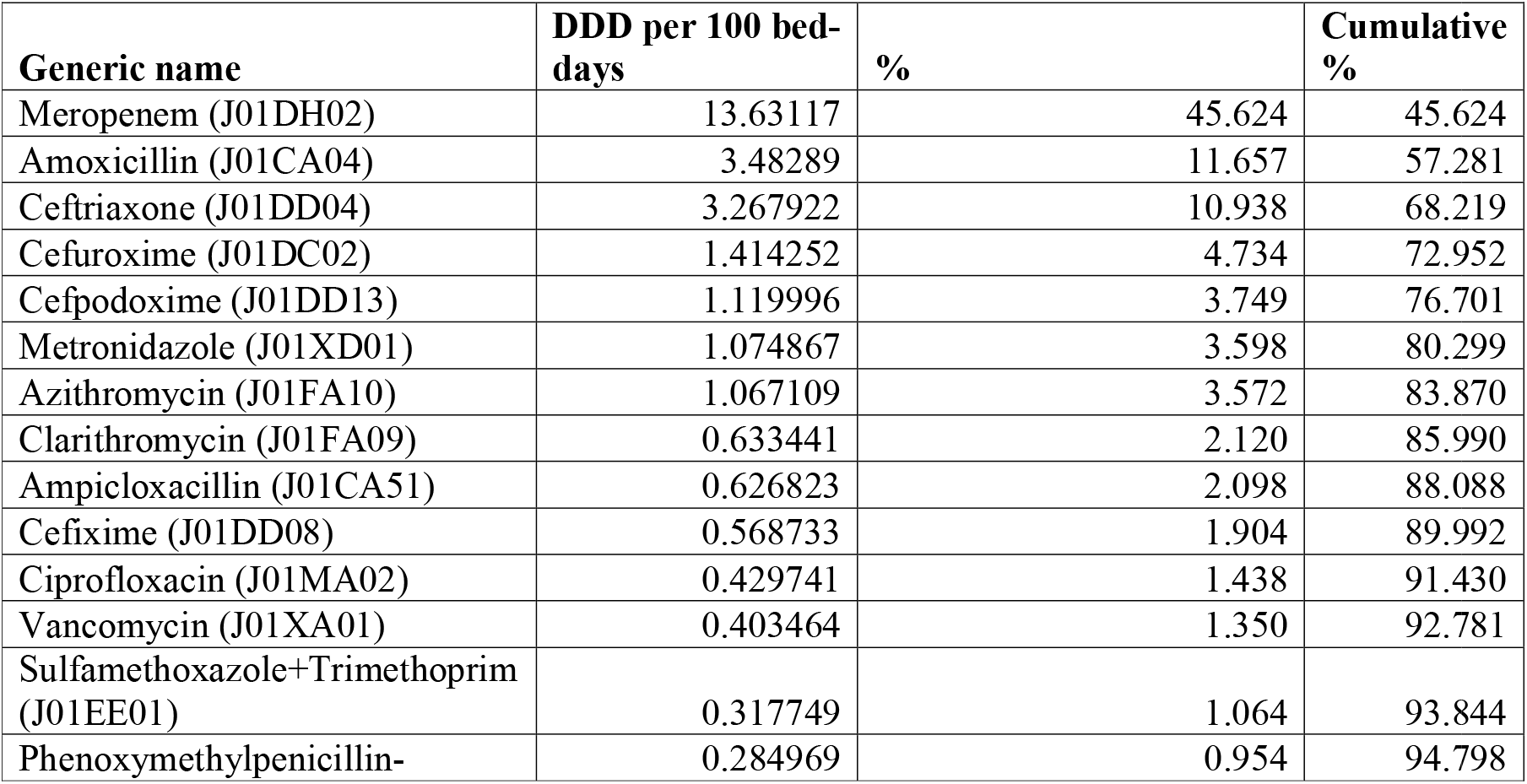
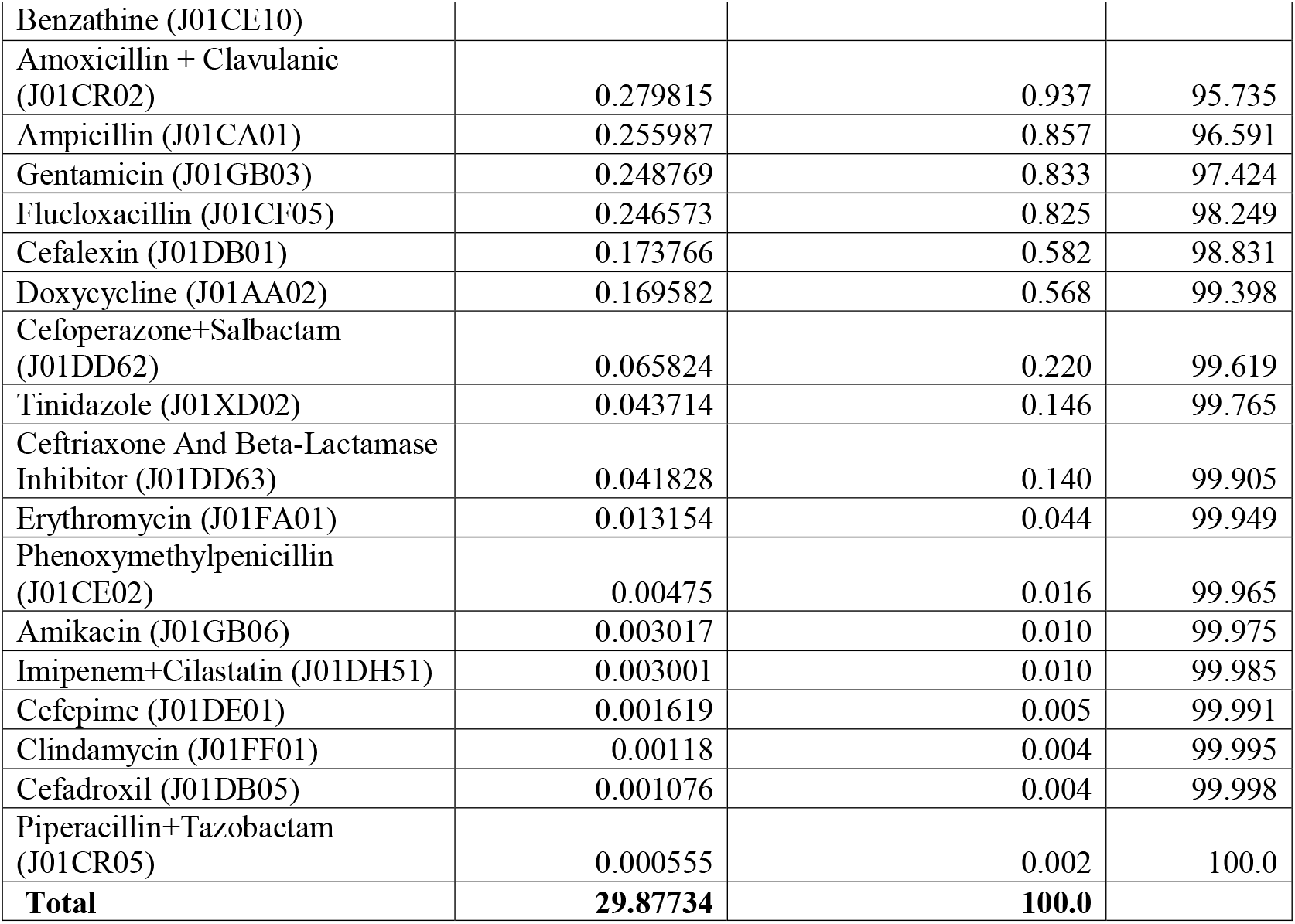
Anatomical Therapeutic Chemical (ATC) Level 5 antibiotic molecules contributing less than 1.5% each to the total inpatient antibiotic use at JKCI from 2016 to 2022.

### Antibiotic use by WHO AWaRE classification among inpatients

Analysis of inpatient antibiotic use by WHO AWaRe classification showed that Reserve group antibiotics contributed the largest share, accounting for 14.04 DDD per 100 bed-days (46.4% of total use), followed by the Watch group at 9.46 DDD per 100 bed-days (31.3%) and the Access group at 6.38 DDD per 100 bed-days (21.1**%**) (Figure 4).

**Figure 4:**
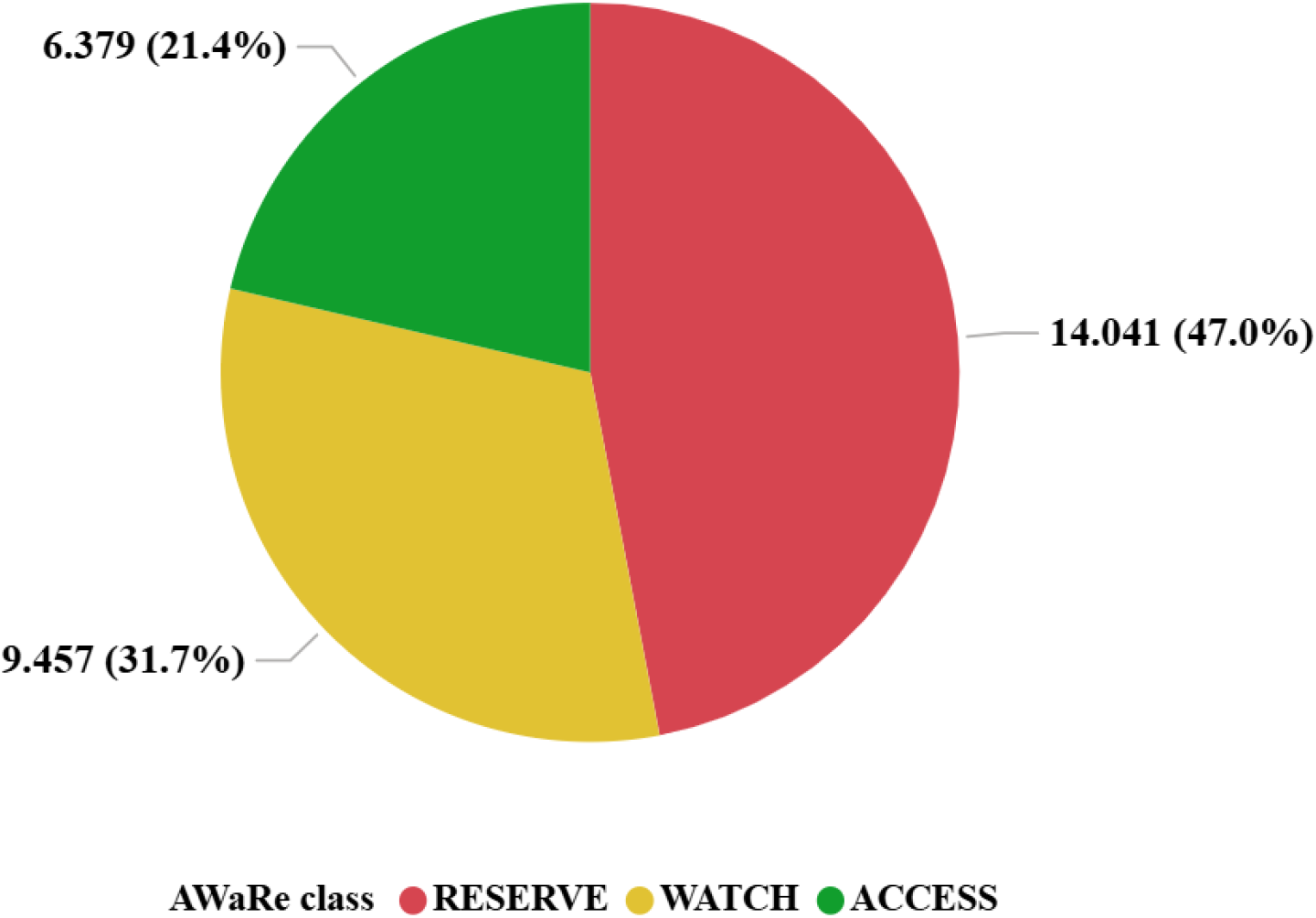
Defined Daily Dose (DDD) proportional contribution of WHO Access, Watch, Reserve (AWaRe)-classified antibiotics among inpatients at JKCI, Tanzania, from 2016 to 2022.

### ARIMA model forecasting future use of antibiotics in inpatients

Observed DDD per 100 bed-days rose from 0.74 in 2016–2017 to 7.79 in 2021–2022, indicating a consistent upward trend in antibiotic use among cardiovascular inpatients. The fitted ARIMA (1,1,0) model projected this increase to continue, with forecasts of 8.50, 8.85, 9.02, and 9.10 for the years 2023 through 2026, respectively. The red forecast line illustrates this growth trajectory, while the green line represents observed data (Figure 5). The prediction intervals remain narrow, indicating relatively stable model confidence in continued escalation.

**Figure 5:**
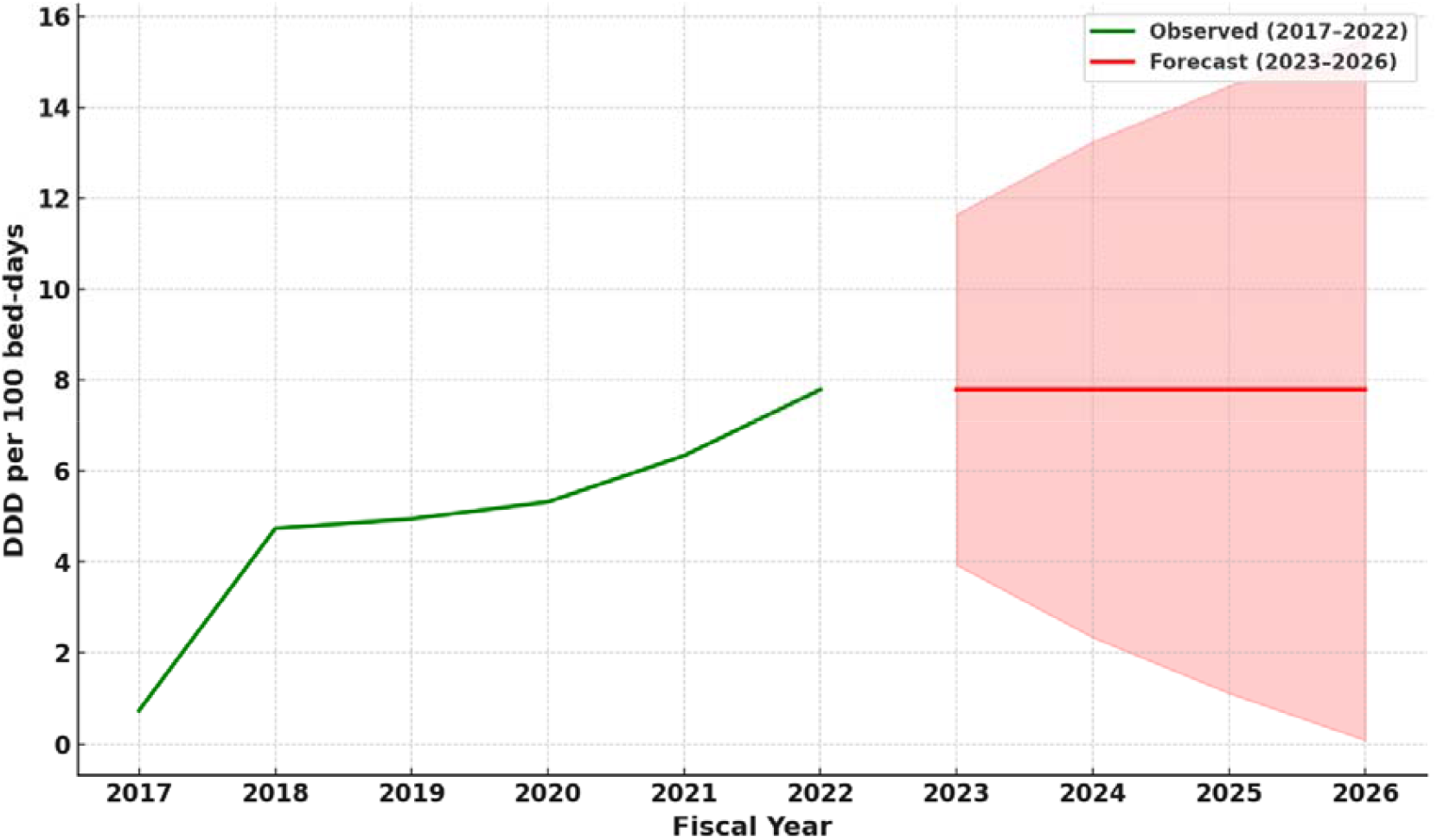
Observed and forecasted Defined Daily Dose (DDD) per 100 bed-days using the Autoregressive Integrated Moving Average (ARIMA) (1,1,0) model, showing trend through 2026.

## 4. Discussion

This study demonstrates that more than half of cardiovascular inpatients at JKCI received systemic antibiotics, underscoring the importance of AMS in specialised cardiac care. JKCI is the only dedicated cardiac referral hospital in Tanzania, and its prescribing patterns therefore provide critical insight into antibiotic use in this unique setting. Overall, antibiotic use was high, dominated by carbapenems, third-generation cephalosporins, and extended-spectrum penicillins. These patterns align with reports from tertiary hospitals in sub-Saharan Africa ^14–16^, but the concentration at JKCI was more pronounced, with meropenem alone contributing nearly half of all inpatient antibiotic use.

Antibiotic exposure varied across patient groups. Children under 10 years and adults over 60 years received the largest share, consistent with other studies that have documented higher use in these age categories ^14–16^. Patients with multi-year admissions also accumulated more exposure, reflecting the complexity of chronic cardiovascular disease management and staged surgical interventions.

Injectable formulations accounted for more than half of the total use, reflecting the severity of illness, high surgical volume, and the need for perioperative prophylaxis. Although tablets were used in step-down therapy, the high reliance on parenteral agents indicates an opportunity for intravenous-to-oral switch protocols in stable patients. Similar interventions have reduced costs and resistance risk in other LMIC hospitals ^7,8^.

At the ATC level 4 classification, prescribing was dominated by carbapenems, third-generation cephalosporins, and extended-spectrum penicillins, with a narrow group of agents exceeding the DU90 threshold. At the level 5 (molecule) level, meropenem alone contributed nearly 46% of total antibiotic use, reflecting both limited therapeutic alternatives and default empirical practices. Comparable patterns have been described in other African referral hospitals, where heavy reliance on ceftriaxone and carbapenems has been linked to rising resistance rates ^22–26^. Beyond concerns about resistance, antibiotic use in cardiovascular patients poses additional risks. For instance, long-term azithromycin use has been linked to increased cardiovascular events in elderly patients ^7^ and disruption of the gut microbiota may elevate the risk of stroke and other CVD outcomes ^8^. These insights underscore the importance of cautious, evidence-based prescribing in this vulnerable population.

The WHO AWaRe framework provides an important benchmark for stewardship. At JKCI, Reserve and Watch antibiotics accounted for nearly 80% of total use, while Access agents made up only one-fifth, well below the WHO recommendation that at least 60% of use be Access ^27^. This imbalance underscores the need for AWaRe-aligned stewardship, including audits, formulary management, and prescriber feedback.

Inappropriate antibiotic use has been widely reported in LMICs ^13^. While empirical treatment is often unavoidable in critical care, persistent reliance on broad-spectrum agents points to gaps in the implementation of AMS. Unlike point prevalence surveys (PPS), which provide snapshots, the DDD per 100 bed-days approach applied here offers a more robust measure for monitoring changes over time ^17,28^.

Forecasting with the ARIMA (1,1,0) model confirmed an upward trajectory in antibiotic use, projecting further increases by 2026. These findings suggest that current prescribing behaviours are strongly shaped by historical patterns and diagnostic limitations, particularly in surgical and intensive care units. However, projections assume the stability of underlying drivers; external shocks such as COVID-19, supply disruptions, or new national policies may alter these trends ^29–31^.

Tanzania’s National Action Plan on AMR (2017–2022) emphasises hospital-level AMS interventions, including real-time monitoring and audit-feedback loops ^32^. The patterns observed at JKCI indicate the need to strengthen these efforts, particularly through regular DU90 audits, formulary restrictions on Reserve antibiotics, and the adoption of IV-to-oral switch protocols. Pairing routine surveillance with predictive tools such as ARIMA can help identify priority areas for intervention and optimise resource allocation.

## 5. Conclusions

Antibiotic use among cardiovascular inpatients at JKCI showed a steady upward trend, with a heavy weighting toward injectable and broad-spectrum agents, and reserve antibiotics contributing a substantial share under the WHO AWaRe framework. As the only specialised cardiac referral hospital in Tanzania, JKCI provides a critical benchmark for understanding antimicrobial use in this high-risk population. Forecasting suggests continued growth, reinforcing the need for hospital-level stewardship strategies to optimise prescribing and preserve the effectiveness of last-line agents.

### Recommendations

Strengthening AMS in tertiary cardiac settings should focus on four priorities. First, implementing intravenous-to-oral switch protocols can reduce hospital stays, costs, and injection-related complications. Second, regular DU90 audits are needed to monitor formulary balance and prevent over-reliance on a few molecules such as meropenem, ceftriaxone, and amoxicillin. Third, stricter oversight of Reserve and Watch antibiotics, supported by diagnostic stewardship, is essential to preserve last-line options. Ultimately, predictive surveillance systems that integrate retrospective DDD monitoring with ARIMA-based forecasting can help identify emerging trends and inform timely interventions. Tailoring prescribing guidance to pediatric and readmitted patients will further enhance rational antibiotic use and safeguard outcomes in this high-risk population.

### Study Limitations

Several limitations should be noted. This analysis was conducted at a single tertiary care centre, which may limit the generalizability of findings to all hospitals or regions. However, JKCI is the only specialised cardiac institute in Tanzania, and its prescribing patterns are likely to influence practice in similar referral settings. The study relied on prescription and dispensing records rather than direct measures of use, which meant that adherence and clinical outcomes could not be assessed. In addition, while DDDs provide a standardised metric, they do not account for pediatric dosing or renal adjustment, which may lead to under- or overestimation of actual use. Future research should expand to multicenter studies and incorporate microbiological and clinical outcome data.

## List of Abbreviations

### Abbreviation Full Term

AMR: Antimicrobial resistance
AMS: Antimicrobial stewardship
ARIMA: AutoRegressive Integrated Moving Average
ATC: Anatomical Therapeutic Chemical classification
AWaRe: Access, Watch, and Reserve classification of antibiotics (WHO)
CHD: Coronary heart disease
CVD: Cardiovascular disease
DDD: Defined Daily Dose
DU90: Drug Utilisation 90%
IV-to-PO: Intravenous-to-oral switch
JKCI: Jakaya Kikwete Cardiac Institute
LMIC: Low- and middle-income country
MUHAS: Muhimbili University of Health and Allied Sciences
NHIF: National Health Insurance Fund
NSSF: National Social Security Fund
PPS: Point Prevalence Survey
SIC: State Insurance Corporation
SMZ: Serikali ya Mapinduzi Zanzibar (Revolutionary Government of Zanzibar)

## Author Contributions

Conceptualisation, J.A.M., R.Z.S. and N.M.; methodology, J.A.N. and R.Z.S.; software, J.A.N.; validation, J.A.N., J.A.M., and R.Z.S.; formal analysis, J.A.N. and R.Z.S.; investigation, J.A.N., H.A.K., and J.M.; resources, M.J. and N.M.; data curation, J.A.N. and J.A.M.; writing—original draft preparation, J.A.N. and R.Z.S.; writing—review and editing, R.Z.S., R.K.M., and J.M.; visualisation, J.A.N.; supervision, R.Z.S. P.K and J.M.; project administration, R.Z.S.; funding acquisition, M.J and R.Z.S. All authors have read and agreed to the published version of the manuscript.

## Funding

This research received no external funding.

## Institutional Review Board Statement

The study was conducted in accordance with the Declaration of Helsinki and approved by the Institutional Review Board of the Muhimbili University of Health and Allied Sciences (MUHAS) with reference number DA.25/111/25/01/2021, and the Jakaya Kikwete Cardiac Institute (JKCI). Ethical clearance was obtained for the secondary use of anonymised hospital prescription records, and no identifiable patient information was accessed.

## Informed Consent Statement

Patient consent was waived for this retrospective study because the data analysed were fully anonymised, routinely collected hospital records, and no identifiable patient information was used. The study posed minimal risk to participants and did not involve direct patient contact.

## Data availability statement

Due to patient confidentiality restrictions, the data cannot be shared publicly. Aggregated datasets may be provided by the corresponding author upon reasonable request.

## Acknowledgments

The authors wish to thank the staff of the Jakaya Kikwete Cardiac Institute (JKCI) for their administrative and technical support during data retrieval and validation.

## Conflicts of interest

All authors report no conflicts of interest relevant to this article.

## Notes

### Competing Interest Statement

The authors have declared no competing interest.

### Author Declarations

The study was approved by the Muhimbili University of Health and Allied Sciences (MUHAS) Research and Ethics Committee under reference number DA.25/111/25/01/2021.

## References

1. Heron M. Deaths: Leading Causes for 2017. Natl Vital Stat Rep. 2019;68(6):1–77. http://www.ncbi.nlm.nih.gov/pubmed/32501203

2. Bochen et al. WHO methods and data sources for country-level causes of death. World Health organisation. 2020. https://www.who.int/docs/default-source/gho-documents/global-health-estimates/ghe2019_cod_methods.pdf

3. Hajar R. Risk Factors for Coronary Artery Disease: Historical Perspectives. Heart Views. 2017;18(3):109–114. doi:10.4103/HEARTVIEWS.HEARTVIEWS_106_17

4. Collaborators G, Roth GA. Global, Regional, and National Burden of Cardiovascular Diseases and Risk Factors in 204 Countries and Territories, 1990-2023. Preprint posted online 2025. doi:10.2139/ssrn.5392535

5. Mensah GA, Fuster V, Murray CJL, et al. Global Burden of Cardiovascular Diseases and Risks, 1990-2022. J Am Coll Cardiol. 2023;82(25):2350–2473. doi:10.1016/j.jacc.2023.11.007

6. Anderson JL, Muhlestein JB. Antibiotic trials for coronary heart disease. Tex Heart Inst J. 2004;31(1):33–38. http://www.ncbi.nlm.nih.gov/pubmed/15061624

7. Zaroff JG, Cheetham TC, Palmetto N, et al. Association of Azithromycin Use With Cardiovascular Mortality. JAMA Netw Open. 2020;3(6):e208199. doi:10.1001/jamanetworkopen.2020.8199

8. Heianza Y, Zheng Y, Ma W, et al. Duration and life-stage of antibiotic use and risk of cardiovascular events in women. Eur Heart J. 2019;40(47):3838–3845. doi:10.1093/eurheartj/ehz231

9. Naimi T, Ringwald P, Besser R, Thompson S. Antimicrobial resistance. Emerg Infect Dis. 2001;7(3 Suppl):548. doi:10.3201/eid0707.010727

10. de Kraker MEA, Stewardson AJ, Harbarth S. Will 10 Million People Die a Year due to Antimicrobial Resistance by 2050? PLoS Med. 2016;13(11):e1002184. doi:10.1371/journal.pmed.1002184

11. Van Boeckel TP, Gandra S, Ashok A, et al. Global antibiotic consumption 2000 to 2010: an analysis of national pharmaceutical sales data. Lancet Infect Dis. 2014;14(8):742–750. doi:10.1016/S1473-3099(14)70780-7

12. Sangeda RZ, Saburi HA, Masatu FC, et al. National Antibiotics Utilisation Trends for Human Use in Tanzania from 2010 to 2016 Inferred from Tanzania Medicines and Medical Devices Authority Importation Data. Antibiotics. 2021;10(10):1249. doi:10.3390/antibiotics10101249

13. Tang KWK, Millar BC, Moore JE. Antimicrobial Resistance (AMR). Br J Biomed Sci. 2023;80. doi:10.3389/bjbs.2023.11387

14. Malo S, José Rabanaque M, Feja C, Jesús Lallana M, Aguilar I, Bjerrum L. High Antibiotic Consumption: A Characterisation of Heavy Users in Spain. Basic Clin Pharmacol Toxicol. 2014;115(3):231–236. doi:10.1111/bcpt.12211

15. Shomuyiwa DO, Lucero_Prisno DE, Manirambona E, et al. Curbing antimicrobial resistance in post_COVID Africa: Challenges, actions and recommendations. Health Sci Rep. 2022;5(5). doi:10.1002/hsr2.771

16. Center for Disease Control and Prevention. Antibiotic Use in the United States, 2020: Progress and Opportunities. 2020. https://www.cdc.gov/antibiotic-use/pdfs/stewardship-report-2020-H.pdf

17. World Health Organisation. Introduction to Drug Utilization Research. 2003. Accessed July 12, 2021. https://www.who.int/publications/i/item/8280820396

18. WHO, Organization World Health. WHO report on surveillance of antibiotic consumption: 2016–2018 early implementation. Geneva: World Health Organisation. World Health Organization. 2018. Accessed September 20, 2023. https://www.who.int/medicines/areas/rational_use/who-amr-amc-report-20181109.pdf?ua=1

19. Mbwasi R, Mapunjo S, Wittenauer R, et al. National Consumption of Antimicrobials in Tanzania: 2017–2019. Front Pharmacol. 2020;11(October):2017–2019. doi:10.3389/fphar.2020.585553

20. Sangeda RZ, William SM, Masatu FC, et al. Antibiotic utilisation patterns in Tanzania: a retrospective longitudinal study comparing pre- and intra-COVID-19 pandemic era using Tanzania Medicines and Medical Devices Authority data. JAC Antimicrob Resist. 2024;6(3). doi:10.1093/jacamr/dlae081

21. Shrestha B, Dixit SM. The Assessment of Drug Use Pattern Using WHO Prescribing Indicators. J Nepal Health Res Counc. 2018;16(3):279–284.

22. Wushouer H, Zhang ZX, Wang JH, et al. Trends and relationship between antimicrobial resistance and antibiotic use in Xinjiang Uyghur Autonomous Region, China: Based on a 3 year surveillance data, 2014–2016. J Infect Public Health. 2018;11(3):339–346. doi:10.1016/J.JIPH.2017.09.021

23. Tadesse BT, Ashley EA, Ongarello S, et al. Antimicrobial resistance in Africa: a systematic review. BMC Infect Dis. 2017;17(1):616. doi:10.1186/s12879-017-2713-1

24. Elton L, Thomason MJ, Tembo J, et al. Antimicrobial resistance preparedness in sub-Saharan African countries. Antimicrob Resist Infect Control. 2020;9(1):145. doi:10.1186/s13756-020-00800-y

25. Horumpende PG, Mshana SE, Mouw EF, Mmbaga BT, Chilongola JO, de Mast Q. Point prevalence survey of antimicrobial use in three hospitals in North-Eastern Tanzania. Antimicrob Resist Infect Control. 2020;9(1):149. doi:10.1186/s13756-020-00809-3

26. Afriyie DK, Sefah IA, Sneddon J, et al. Antimicrobial point prevalence surveys in two Ghanaian hospitals: opportunities for antimicrobial stewardship. JAC Antimicrob Resist. 2020;2(1). doi:10.1093/jacamr/dlaa001

27. WHO. 2021 AWaRe classification. Accessed October 9, 2023. https://www.who.int/publications/i/item/2021-aware-classification

28. Ansari F, Erntell M, Goossens H, Davey P. The European Surveillance of Antimicrobial Consumption (ESAC) Point□Prevalence Survey of Antibacterial Use in 20 European Hospitals in 2006. Clinical Infectious Diseases. 2009;49(10):1496–1504. doi:10.1086/644617

29. Box GEP., Jenkins GM., Reinsel GC., Ljung GM. Time Series Analysis□: Forecasting and Control. John Wiley & Sons, Inc.; 2016.

30. Murray CJL, Ikuta KS, Sharara F, et al. Global burden of bacterial antimicrobial resistance in 2019: a systematic analysis. The Lancet. 2022;399(10325):629–655. doi:10.1016/S0140-6736(21)02724-0

31. Lanckohr C, Bracht H. Antimicrobial stewardship. Curr Opin Crit Care. 2022;28(5):551–556. doi:10.1097/MCC.0000000000000967

32. United Republic of Tanzania. The National Action Plan on Antimicrobial Resistance 2017 - 2022. 2017. Accessed November 6, 2019. https://www.afro.who.int/publications/national-action-plan-antimicrobial-resistance-2017-2022

